# Pre-diagnostic lipid metabolites are enriched in men who develop advanced prostate cancer: a nested case-control study

**DOI:** 10.64898/2026.03.12.26348193

**Authors:** Rebecca E. Graff, Harriett Fuller, Kathryn M. Wilson, Barbra A. Dickerman, June M. Chan, Philip W. Kantoff, Xiaoshuang Feng, Clary B. Clish, Matthew G. Vander Heiden, Burcu F. Darst, Ericka M. Ebot, Lorelei A. Mucci

## Abstract

Few studies with pre-diagnostic samples have estimated associations between circulating metabolites and risk of advanced prostate cancer. We performed untargeted metabolomic profiling of pre-diagnostic blood samples from 212 advanced prostate cancer cases (stage ≥T3b or lethal during follow-up) and 212 matched controls from the Health Professionals Follow-up Study. 243 metabolites were assayed using liquid chromatography-tandem mass spectrometry (Broad Institute) and met quality control standards. We used multivariable conditional logistic regression to generate odds ratios (OR) and 95% confidence intervals (95%CI) for associations between individual metabolites and risk of advanced prostate cancer, and conducted metabolite set enrichment tests to identify metabolite classes enriched in advanced prostate cancer. Subgroup analyses were conducted by body mass index (BMI) and time between blood draw and diagnosis. Levels of 16 lipid species were nominally associated with advanced prostate cancer at *p*<0.05, though none were statistically significant after multiple testing correction. The strongest signals were for C56:1 triacylglycerol (TAG; OR: 1.34, 95%CI: 1.07-1.67) and C38:4 diacylglycerol (DAG; OR: 1.27, 95%CI: 1.04-1.55). Enrichment analyses revealed six metabolite classes associated with advanced prostate cancer after multiple testing adjustment, the top four of which were DAGs and TAGs: DAGs overall (*P*=3.4E-07), unsaturated DAGs (*P*=5.9E-07), unsaturated TAGs (*P*=2.3E-06), and TAGs overall (*P*=2.4E-06). 43 metabolites were nominally associated with advanced prostate cancer among individuals with BMI <25 kg/m^2^; only three demonstrated nominal associations in individuals with BMI ≥25 kg/m^2^. These findings suggest associations between circulating pre-diagnostic lipid levels and aggressive prostate cancer risk, particularly in lean individuals.

## INTRODUCTION

Prostate cancer is the second leading cause of cancer death among men in the United States.^1^ Identifying risk factors for *advanced* disease holds promise for prevention of and reductions in fatal prostate cancer. One possible set of such factors relates to metabolic health. Adiposity, and particularly central obesity, is a risk factor for advanced prostate cancer,^2-5^ prostate tumors rely on *de novo* lipogenesis for growth,^6^ and blood-based biomarkers of lipid-related metabolism are associated with disease progression.^7^

Early discovery-based epidemiologic studies of circulating metabolites have shed light on specific metabolites in the etiology of prostate cancer.^8^ However, a limited number of study populations have collected blood samples years prior to diagnosis, such that they were well positioned and powered to prospectively interrogate circulating metabolite levels associated with future advanced disease risk.^9^ Additional studies are needed to validate prior results and identify further metabolites and metabolite classes associated with advanced prostate cancer.

We conducted a prospective investigation nested in the Health Professionals Follow-up Study (HPFS) that compared pre-diagnostic metabolite levels between men with advanced prostate cancer and matched controls. Given the effect of obesity on the metabolome,^10^ we evaluated whether body mass index (BMI) modifies associations between metabolites and advanced prostate cancer risk. We additionally examined timing between blood draw and diagnosis to shed light on potential biomarkers for early detection (little time between blood draw and diagnosis) and etiology (longer time between blood draw and diagnosis). Evaluation of metabolite classes and network analysis further aided in identifying molecular pathways of metabolic alterations in the development of advanced prostate cancer.

## MATERIALS AND METHODS

### Study Population

The HPFS is an ongoing prospective cohort of 51,529 male health professionals age 40-75 at enrollment in 1986.^11^ Participants completed a baseline questionnaire concerning medical history and lifestyle factors, and have since completed follow-up questionnaires every two years thereafter. This study was nested within the blood sub-cohort of 18,159 participants free of prostate cancer diagnosis at blood collection in 1993-1995.^12^ Blood samples were centrifuged to collect plasma that was then stored in liquid nitrogen freezers (<130°C). Participants reported the timing of blood collection and hours fasted prior to blood collection on a questionnaire returned with the samples.

We undertook a nested case-control study of metabolomics and advanced prostate cancer. Incident prostate cancer cases were confirmed via medical records or pathology reports. Study investigators reviewed records and prostate cancer-specific follow-up questionnaires to abstract information about tumor stage (clinical and/or pathological), Gleason score, and development of metastases over time. Deaths were confirmed by an endpoints committee using medical records and death certificates as well as the National Death Index to determine cause of death, including from prostate cancer. The 212 advanced prostate cancer cases included in this nested case-control study were comprised of all men diagnosed between the time of blood draw and September 2010 with an available plasma sample who: 1) died due to prostate cancer (n=111); 2) developed metastases during follow-up (n=33) but had not (yet) died due to prostate cancer; or 3) were stage T3b, T4, N1, and/or M1 at diagnosis (n=68) and had not (yet) demonstrated evidence of prostate cancer death or metastases. Since initial matching, an additional six of the metastatic cases and 10 of the high-stage cases died due to prostate cancer, and an additional high-stage case developed metastases (but had not died due to prostate cancer as of March 6, 2024).

For each case, one control was selected who was cancer-free at time of the case’s diagnosis, resulting in 212 included controls. Individual matching criteria were age (±1 year), completed prostate-specific antigen testing between January 1992 and blood draw (yes, no), and blood collection time of day (midnight-9am, 9am-noon, noon-4pm, 4pm-midnight), season (winter, spring, summer, fall), and year (exact). Since matching, 14 of the controls were diagnosed with prostate cancer, including three who would have met the criteria for advanced prostate cancer.

### Metabolomic Profiling

Plasma samples for the 424 participants and 10% quality control replicate samples were analyzed at the Broad Institute (Cambridge, MA) as described previously.^12, 13^ In brief, metabolites were quantified using using two liquid chromatography-tandem mass spectrometry methods that measured: 1) amines and polar metabolites that ionize in the positive ion mode and 2) polar and non-polar lipids. The lab was blinded to case-control status, and matched pairs were run on adjacent wells. Out of the 308 metabolites assessed, this study describes analyses of 243 known metabolites with covariate of variation ≤25% and intraclass correlation coefficient ≥0.4 (**Supplementary Table S1**). Peaks were log(10)-transformed to improve normality and standardized by dividing peaks by the standard deviation in the population. To identify biologically meaningful metabolite pathways, the Broad Institute assigned metabolites to the following classes defined based on chemical taxonomy: 28 amines, 6 amino acid derivatives, 25 amino acids, 177 lipids and lipid metabolites, 3 purines, pyrimidines, and derivatives, 1 sphingolipid, and 3 ‘other’ metabolites.

### Assessment of BMI

BMI (kg/m^2^) at blood draw was determined by weight reported on the questionnaire immediately preceding blood collection and height reported at baseline in 1986. In a validation study, self-reported weights were strongly correlated with technician-measurements (Pearson *r*=0.97).^14^ Men with BMI <25 kg/m^2^ were considered lean, and men with BMI ≥25 kg/m^2^ were considered overweight/obese.

### Statistical Analysis

Conditional logistic regression was implemented to estimate odds ratios (OR) and 95% confidence intervals (CI) for associations between individual metabolites and risk of advanced prostate cancer. These models inherently adjusted for matching factors and were additionally adjusted for fasting status (<4 hours, 4-7 hours, 8-11 hours, ≥12 hours, missing). Models further adjusting for BMI (kg/m^2^, continuous) and, separately, cholesterol levels (mg/dL, continuous) were also fit, restricting to individuals without missingness (n=410 for BMI-adjusted, n=310 for cholesterol-adjusted).

To identify metabolite classes enriched in advanced prostate cancer risk, we implemented geneSetTest from R’s limma package.^15^ These tests used Wilcoxon rank sum tests to evaluate whether the rank of each metabolite class, determined by *p*-values for associations between individual metabolites and advanced prostate cancer, were higher than expected by chance. We assigned positive or negative directionality to the results of the enrichment analyses by qualitatively assessing the direction of associations for the metabolites within each class.

To examine latency periods, we evaluated associations for cases below and, separately, above the median time between blood draw and prostate cancer diagnosis or index date for controls (5.5 years).. This analysis distinguished markers of early detection (below the median) versus etiology (above the median). To determine whether associations of individual metabolites or metabolite classes were distinct in participant subgroups defined by clinical characteristics, we also evaluated associations for low Gleason score (≤3+4) and higher Gleason score (≥4+3) advanced prostate cancer cases separately. Given broad effects of obesity on the metabolic milieu, we additionally stratified by BMI at blood draw (<25 kg/m^2^, ≥25 kg/m^2^) and calculated a Pearson correlation between each metabolite and BMI in the study population. For metabolites that were nominally statistically significant in one stratum in a pair (defined by time between blood draw and diagnosis, Gleason score, or BMI) but not the other, we assessed the Q-statistic for heterogeneity across strata via random effects meta-analysis.

Finally, we used weighted correlation network analysis (WGCNA) to identify networks of highly correlated metabolites. In brief, the method entailed calculating pairwise Pearson correlations^16^ and constructing a weighted adjacency matrix using a soft thresholding power of six.^17^ Average linkage hierarchical clustering was used to construct the topological overlap matrix’s clustering tree structure to identify modules of densely interconnected metabolites.^18^ Principal component analyses were then conducted for each module. The module’s first principal component, which captured most variation in metabolite values, was tested for associations with advanced prostate cancer using conditional logistic regression, as above.

Analyses were conducted using SAS statistical software, version 9.3 (SAS Institute, Cary, NC) or R version 3.6.3.

## RESULTS

Characteristics of the study population are presented in **Table 1**. As expected, the distributions of matching factors were comparable between cases and controls. Cases and controls had similar BMI and cholesterol levels, though cases were slightly more likely to have fasted before blood draw. Among cases, the median time between blood draw and cancer diagnosis was 5.5 years (interquartile range: 3.0, 9.0). Three-quarters (75%) of the advanced prostate cancers were initially diagnosed with localized stage disease, and 22% had Gleason score ≤6. Most cases (62%) were initially treated with radical prostatectomy or radiation.

**Table 1.** Characteristics of advanced prostate cancer cases^a^ and matched controls in the Health Professionals Follow-up Study, median (interquartile range) or n (%)

|  | Advanced Cases<br>n = 212 | Controls<br>n = 212 |
| --- | --- | --- |
| **Characteristics at Blood Draw** |  |  |
| Age at Blood Draw<sup>b</sup>, years | 67 (60, 72) | 66 (60, 71) |
| Self-reported Race |  |  |
| Asian<sup>c</sup> | 0 (0.0%) | 1 (0.5%) |
| Black<sup>c</sup> | 1 (0.5%) | 1 (0.5%) |
| White<sup>c</sup> | 204 (99%) | 195 (97%) |
| Other<sup>c</sup> | 1 (0.5%) | 5 (2.5%) |
| Missing, n | 6 | 10 |
| Ever PSA Test<sup>b</sup> | 132 (62%) | 128 (60%) |
| Time of Day<sup>b</sup> |  |  |
| 12 am to before 9 am<sup>c</sup> | 57 (28%) | 58 (29%) |
| 9 am to before 12 pm<sup>c</sup> | 98 (48%) | 98 (49%) |
| 12 pm to before 4 pm<sup>c</sup> | 41 (20%) | 39 (19%) |
| 4pm to before 12am<sup>c</sup> | 8 (4%) | 6 (3%) |
| Missing, n | 8 | 11 |
| Season of Blood Draw<sup>b</sup> |  |  |
| Winter | 28 (13%) | 25 (12%) |
| Spring | 42 (20%) | 49 (23%) |
| Summer | 77 (36%) | 77 (36%) |
| Fall | 65 (31%) | 61 (29%) |
| Year of Blood Draw<sup>b</sup> |  |  |
| 1993 | 99 (47%) | 100 (47%) |
| 1994 | 106 (50%) | 106 (50%) |
| 1995 | 7 (3%) | 6 (3%) |
| ≥8 Hours of Fasting<sup>c</sup> | 141 (73%) | 132 (68%) |
| Missing, n | 18 | 19 |
| BMI, kg/m<sup>2</sup><sup>c</sup> | 25 (23, 28) | 26 (23, 28) |
| Missing, n | 10 | 4 |
| Total Blood Cholesterol, mg/dL<sup>c</sup> | 196 (178, 220) | 197 (177, 217) |
| Missing, n | 56 | 58 |
| **Case Characteristics** |  |  |
| Time from Blood Draw to Diagnosis, years | 5.5 (2.5, 9.2) | - |
| Fatal Prostate Cancer | 111 (52%) | - |
| Stage at Diagnosis |  |  |
| T1<sup>c</sup> | 30 (15%) | - |
| T2<sup>c</sup> | 45 (23%) | - |
| T3 or T3a<sup>c</sup> | 11 (6%) | - |
| T3b or T4<sup>c</sup> | 62 (31%) | - |
| N1/M0<sup>c</sup> | 18 (9%) | - |
| M1<sup>c</sup> | 32 (16%) | - |
| Missing, n | 14 |  |
| Gleason Score at Diagnosis<sup>c</sup> |  |  |
| ≤6<sup>c</sup> | 41 (23%) | - |
| 3+4<sup>c</sup> | 37 (20%) | - |
| 4+3<sup>c</sup> | 35 (19%) | - |
| 8-10<sup>c</sup> | 69 (38%) | - |
| Missing, n | 30 |  |
| Primary Treatment |  |  |
| Radical Prostatectomy<sup>c</sup> | 83 (43%) | - |
| Radiation<sup>c</sup> | 49 (25%) | - |
| Hormones<sup>c</sup> | 32 (17%) | - |
| Active Surveillance/None<sup>c</sup> | 18 (9%) | - |
| Other or Unknown<sup>c</sup> | 11 (7%) | - |
| Missing, n | 19 | - |

Abbreviations: BMI, body mass index; PSA, prostate-specific antigen
Note: percentages may not add up to 100% due to rounding
<sup>a</sup> Individuals who 1) died due to prostate cancer (n = 111); 2) developed metastases during follow-up (n = 33) but had not (yet) died due to prostate cancer; or 3) were stage T3b, T4, N1 and/or M1 at diagnosis (n = 68) and had not (yet) demonstrated evidence of prostate cancer death or metastases
<sup>b</sup> Matching factor
<sup>c</sup> Among individuals with data

Based on a false discovery rate threshold of *q*=0.1 (to account for multiple testing), none of the 243 metabolites were statistically significantly associated with advanced prostate cancer. With nominal significance (*p*<0.05), 16 lipid metabolites were associated with advanced disease risk (**Figure 1**, **Table 2**, **Supplementary Table S2**), including positive associations for 10 unsaturated glycerolipids (six triacylglycerols and four diacylglycerols), with the strongest signals for C56:1 triacylglycerol (OR: 1.34, 95% CI: 1.07-1.67) and C38:4 diacylglycerol (OR: 1.27, 95% CI: 1.04-1.55). Out of the remaining associated metabolites, three acylcarnitines and three plasmalogens were nominally inversely associated with advanced prostate cancer. In BMI-adjusted models, the same acylcarnitines and plasmalogens remained nominally inversely associated with advanced prostate cancer, whereas three of the 10 glycerolipids remained positively associated (**Figure 1**, **Supplementary Table S2**). In cholesterol-adjusted models, there were nominal positive associations for 16 glycerolipids, the amino acid isoleucine, and ceramide (d18:1/22:0). Cholesterol-adjusted models demonstrated inverse associations for C9 carnitine and C36:1 phosphatidylserine plasmalogen (**Figure 1**, **Supplementary Table S2**).

**Figure 1.**
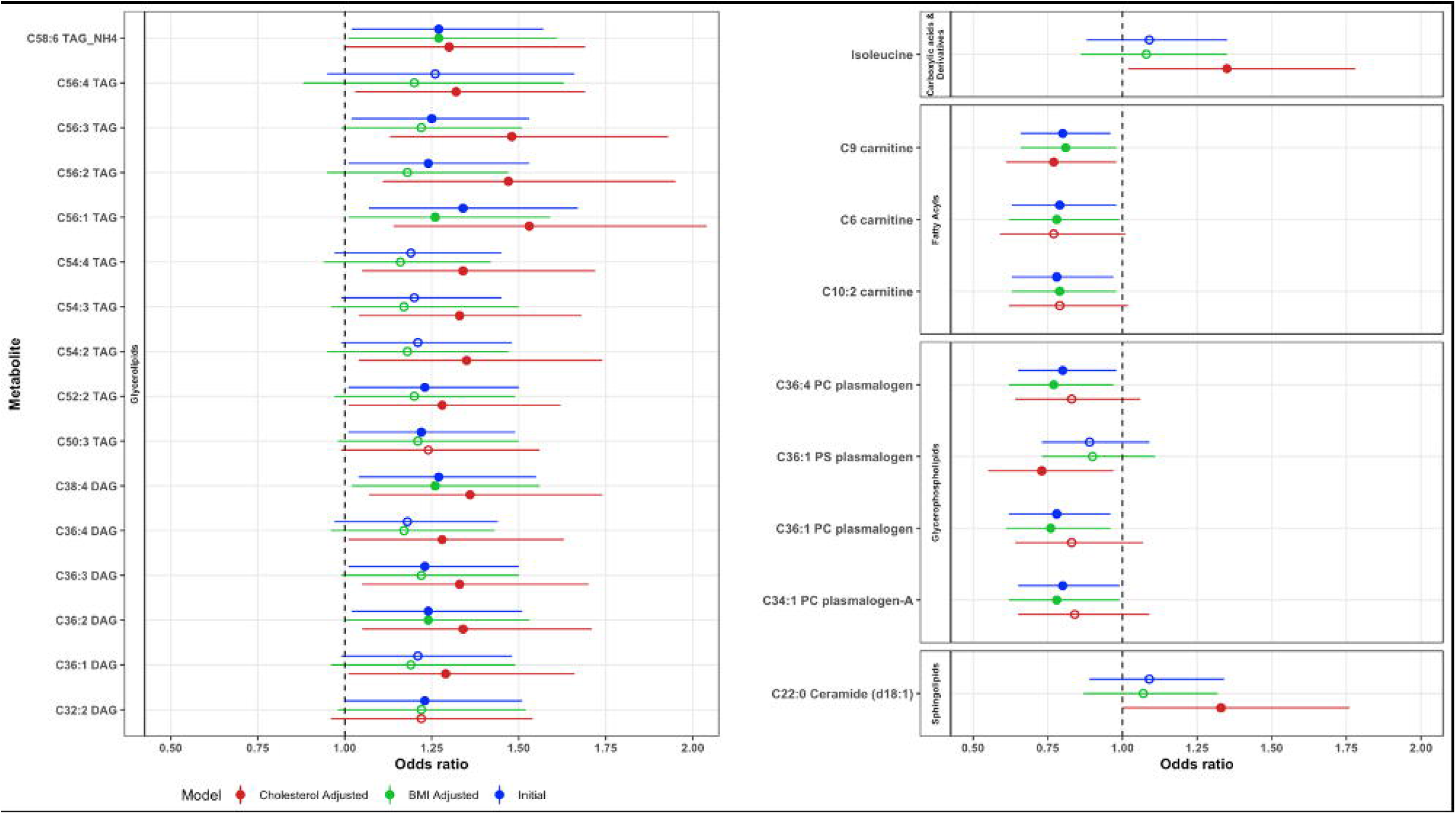
Forest plot highlighting metabolites nominally associated with advanced prostate cancer with *p*<0.05 in at least one analysis. Solid circles indicate *p*<0.05 while open circles indicate *p*≥0.05.

**Table 2.** Odds ratios and 95% confidence intervals for plasma metabolites (per standard deviation) nominally significantly associated with the risk of advanced prostate cancer in the Health Professionals Follow-up Study, *p*<0.05

| HMDB ID<sup>a</sup> | Metabolite Name | OR (95% CI) | *p*-value | *q*-value |
| --- | --- | --- | --- | --- |
| **Unsaturated Diacylglycerols** |  |  |  |  |
| HMDB07170 | C38:4 DAG | 1.27 (1.04, 1.55) | 0.019 | 0.56 |
| HMDB07218 | C36:2 DAG | 1.24 (1.02, 1.51) | 0.030 | 0.56 |
| HMDB07219 | C36:3 DAG | 1.23 (1.01, 1.50) | 0.039 | 0.56 |
| HMDB07128 | C32:2 DAG | 1.23 (1.00, 1.51) | 0.046 | 0.56 |
| **Unsaturated Triacylglycerols** |  |  |  |  |
| HMDB05396 | C56:1 TAG | 1.34 (1.07, 1.67) | 0.0098 | 0.56 |
| HMDB05410 | C56:3 TAG | 1.25 (1.02, 1.53) | 0.029 | 0.56 |
| X952_8313mz_12_19 | C58:6 TAG_NH4 | 1.27 (1.02, 1.57) | 0.029 | 0.56 |
| HMDB05369 | C52:2 TAG | 1.23 (1.01, 1.50) | 0.035 | 0.56 |
| HMDB05404 | C56:2 TAG | 1.24 (1.01, 1.53) | 0.040 | 0.56 |
| HMDB05433 | C50:3 TAG | 1.22 (1.01, 1.49) | 0.043 | 0.56 |
| **Acylcarnitines** |  |  |  |  |
| HMDB13288 | C9 carnitine | 0.80 (0.66, 0.96) | 0.020 | 0.56 |
| HMDB13325 | C10:2 carnitine | 0.78 (0.63, 0.97) | 0.023 | 0.56 |
| HMDB00705 | C6 carnitine | 0.79 (0.63, 0.98) | 0.032 | 0.56 |
| **Plasmalogens** |  |  |  |  |
| HMDB11241 | C36:1 PC plasmalogen | 0.78 (0.62, 0.96) | 0.021 | 0.56 |
| HMDB11310 | C36:4 PC plasmalogen | 0.80 (0.65, 0.98) | 0.034 | 0.56 |
| HMDB11208 | C34:1 PC plasmalogen-A | 0.80 (0.65, 0.99) | 0.043 | 0.56 |
| **Models Adjusted for Body Mass Index** |  |  |  |  |
| **Unsaturated Diacylglycerols** |  |  |  |  |
| HMDB07170 | C38:4 DAG | 1.26 (1.02, 1.56) | 0.029 | 0.68 |
| **Unsaturated Triacylglycerols** |  |  |  |  |
| X952_8313mz_12_19 | C58:6 TAG_NH4 | 1.27 (1.01, 1.61) | 0.041 | 0.68 |
| HMDB05396 | C56:1 TAG | 1.26 (1.01, 1.59) | 0.045 | 0.68 |
| **Acylcarnitines** |  |  |  |  |
| HMDB13288 | C9 carnitine | 0.81 (0.66, 0.98) | 0.034 | 0.68 |
| HMDB13325 | C10:2 carnitine | 0.79 (0.63, 0.98) | 0.035 | 0.68 |
| HMDB00705 | C6 carnitine | 0.79 (0.63, 1.00) | 0.049 | 0.68 |
| **Plasmalogens** |  |  |  |  |
| HMDB11241 | C36:1 PC plasmalogen | 0.76 (0.61, 0.96) | 0.020 | 0.68 |
| HMDB11310 | C36:4 PC plasmalogen | 0.77 (0.62, 0.97) | 0.026 | 0.68 |
| HMDB11208 | C34:1 PC plasmalogen-A | 0.78 (0.62, 0.99) | 0.037 | 0.68 |
| **Models Adjusted for Cholesterol Levels** |  |  |  |  |
| **Unsaturated Diacylglycerols** |  |  |  |  |
| HMDB07170 | C38:4 DAG | 1.36 (1.07, 1.74) | 0.013 | 0.50 |
| HMDB07218 | C36:2 DAG | 1.34 (1.05, 1.71) | 0.018 | 0.50 |
| HMDB07219 | C36:3 DAG | 1.33 (1.05, 1.70) | 0.019 | 0.50 |
| HMDB07248 | C36:4 DAG | 1.28 (1.01, 1.63) | 0.041 | 0.50 |
| HMDB07216 | C36:1 DAG | 1.29 (1.01, 1.66) | 0.044 | 0.50 |
| **Unsaturated Triacylglycerols** |  |  |  |  |
| HMDB05410 | C56:3 TAG | 1.48 (1.13, 1.93) | 0.004 | 0.50 |
| HMDB05396 | C56:1 TAG | 1.53 (1.14, 2.04) | 0.004 | 0.50 |
| HMDB05404 | C56:2 TAG | 1.47 (1.11, 1.95) | 0.008 | 0.50 |
| HMDB05370 | C54:4 TAG | 1.34 (1.05, 1.72) | 0.018 | 0.50 |
| HMDB05405 | C54:3 TAG | 1.33 (1.04, 1.68) | 0.021 | 0.50 |
| HMDB05403 | C54:2 TAG | 1.35 (1.04, 1.74) | 0.023 | 0.50 |
| HMDB05398 | C56:4 TAG | 1.32 (1.03, 1.69) | 0.028 | 0.50 |
| HMDB05384 | C52:3 TAG | 1.30 (1.02, 1.64) | 0.032 | 0.50 |
| HMDB05363 | C52:4 TAG | 1.30 (1.02, 1.67) | 0.034 | 0.50 |
| HMDB05369 | C52:2 TAG | 1.28 (1.01, 1.62) | 0.038 | 0.50 |
| X952_8313mz_12_19 | C58:6 TAG_NH4 | 1.30 (1.00, 1.69) | 0.048 | 0.50 |
| **Acylcarnitines** |  |  |  |  |
| HMDB13288 | C9 carnitine | 0.77 (0.61, 0.98) | 0.031 | 0.50 |
| **Plasmalogens** |  |  |  |  |
| X776_5788mz_8_04 | C36:1 PS plasmalogen | 0.73 (0.55, 0.97) | 0.027 | 0.50 |
| **Amino Acids** |  |  |  |  |
| HMDB00172 | isoleucine | 1.35 (1.02, 1.78) | 0.034 | 0.50 |
| **Ceramides** |  |  |  |  |
| HMDB04952 | C22:0 Ceramide (d18:1) | 1.33 (1.00, 1.76) | 0.048 | 0.50 |

<sup>a</sup> If available

Enrichment analyses revealed six metabolite classes associated with advanced prostate cancer (*q*<0.1, **Table 3**). Diacylglycerols were most strongly positively associated (*q*=6.5×10^-6^), and particularly unsaturated diacylglycerols (*q*=6.5×10^-6^). (As stated in the Methods, directionality was inferred by qualitatively assessing the direction of associations for individual diacylglycerols.) Similarly, both unsaturated triacylglycerols (*q*=1.3×10^-5^) and overall triacylglycerols (*q*=1.3×10^-5^) were significantly positively enriched. Amines (*q*=0.003) and, specifically, acylcarnitines (*q*=3.4×10^-4^) were significantly negatively enriched. These six classes were enriched after adjusting metabolite associations for BMI and, separately, cholesterol levels (**Supplementary Table S3**). BMI-adjusted analyses also found plasmalogens (*q*=0.03) to be negatively enriched with advanced disease, while cholesterol-adjusted analyses rendered ceramides positively enriched (*q*=0.05).

**Table 3.** Associations between (non-mutually exclusive) plasma metabolite classes and risk of advanced prostate cancer in the Health Professionals Follow-up Study, ranked by *q*-value

| **Metabolite Class** | ***p*-value** | ***q*-value** |
| --- | --- | --- |
| Diacylglycerols | $3.4 \times 10^{-7}$ | $6.5 \times 10^{-6}$ |
| Unsaturated Diacylglycerols | $5.9 \times 10^{-7}$ | $6.5 \times 10^{-6}$ |
| Unsaturated Triacylglycerols | $2.3 \times 10^{-6}$ | $1.3 \times 10^{-5}$ |
| Triacylglycerols | $2.4 \times 10^{-6}$ | $1.3 \times 10^{-5}$ |
| Acylcarnitines | $7.8 \times 10^{-5}$ | $3.4 \times 10^{-4}$ |
| Amines | 0.001 | 0.003 |
| Saturated Diacylglycerols | 0.07 | 0.23 |
| Purines, Pyrimidines, and Derivatives | 0.09 | 0.25 |
| Plasmalogens | 0.17 | 0.42 |
| Amino Acid Derivatives | 0.19 | 0.42 |
| Ceramides | 0.25 | 0.50 |
| Saturated Triacylglycerols | 0.32 | 0.58 |
| Lipids and Lipid Metabolites | 0.39 | 0.65 |
| Phosphatidylserines | 0.65 | 1.00 |
| Other | 0.86 | 1.00 |
| Phosphatidylethanolamines | 0.86 | 1.00 |
| Sphingomyelins | 0.96 | 1.00 |
| Phosphatidylcholines | 0.97 | 1.00 |
| Lysophosphatidylcholines | 1.00 | 1.00 |
| Cholesterol Esters | 1.00 | 1.00 |
| Lysophosphatidylethanolamines | 1.00 | 1.00 |
| Amino Acids | 1.00 | 1.00 |

Analyses of cases diagnosed <5.5 years since blood draw (i.e., early cases) yielded 13 metabolites nominally associated with advanced prostate cancer, although none met a *q*=0.1 threshold (**Supplementary Table S4**). Among early cases, the metabolite classes were statistically significantly positively enriched for overall (*q*=1.3×10^-5^) and unsaturated diacylglycerols (*q*=2.9×10^-5^), overall (*q*=1.3×10^-5^) and unsaturated triacylglycerols (*q*=2.9×10^-5^), and lipid and lipid metabolites (*q*=0.001) (**Supplementary Table S5**). For later cases (≥5.5 years since blood draw), there were six metabolites nominally associated with advanced prostate cancer, with no associated metabolites shared with the early cases (**Supplementary Table S4**). Classes for the later cases were also distinct, with associations most strongly enriched for acylcarnitines (*q*=1.0×10^-4^) and amines (*p*=1.3×10^-4^), both negatively (**Supplementary Table S5**). Diacylglycerols and unsaturated diacylglycerols were significantly positively enriched in analyses of early and later cases.

While all cases met the definition of advanced stage prostate cancer, 78 were diagnosed with Gleason score ≤3+4 tumors (i.e, lower grade) and 104 were diagnosed with Gleason score ≥4+3 (i.e, higher grade) tumors. Although power was limited to detect heterogeneity, C56:3 triacylglycerol and C38:4 diacylglycerol were nominally associated with lower grade advanced prostate cancer risk (**Supplementary Table S4**). Lower grade advanced tumors showed statistically significant positive enrichment for lipids and lipid metabolites (*q*=4.6×10^-6^), diacylglycerols (*q*=9.1×10^-4^), and unsaturated diacylglycerols (*q*=9.1×10^-4^), and significant negative enrichment for lysophosphatidylcholines (*q*=0.003). Of the seven metabolites nominally associated with higher grade advanced prostate cancer, C36:4 phosphatidylethanolamine plasmalogen demonstrated nominal evidence of heterogeneity across grade strata (*p*=0.03). Higher grade advanced tumors had associations statistically significantly negatively enriched for plasmalogens (*q*=0.003), acylcarnitines (*q*=0.005), and amines (*q*=0.03) (**Supplementary Table S5**).

BMI was statistically significantly (*q*<0.10) correlated with 135 (56%) of the studied metabolites (**Supplementary Table S6**). Nearly one-fifth (n=48) of all 243 metabolites were nominally associated with advanced prostate cancer among lean individuals, though none demonstrated statistical evidence of heterogeneity. Only three metabolites demonstrated nominal associations in individuals who were overweight/obese (**Supplementary Table S4**), one of which (dimethylglycine) was nominally heterogeneous across strata (*p*=0.02). Among lean individuals, the metabolite classes positively enriched for advanced prostate cancer included overall (*q*=7.5×10^-7^) and unsaturated triacylglycerols (*q*=3.1×10^-6^), overall (*q*=2.8×10^-6^) and unsaturated diacylglycerols (*q*=8.7×10^-6^), and lipid and lipid metabolites (*q*=0.01). Plasmalogens were negatively enriched (*q*=0.005). Among individuals who were overweight/obese, metabolites were statistically significantly negatively enriched for acylcarnitines (*q*=4.7×10^-7^) and amines (*p*=1.6×10^-5^) (**Supplementary Table S5**).

WGCNA identified five metabolite modules (**Supplementary Figure S1, Supplementary Table S7**). Although modules were not statistically significant, module associations with advanced prostate cancer aligned with the other findings; a module largely comprised of acylcarnitines (green) was associated with lower advanced prostate cancer risk, while a module comprised of unsaturated triacylglycerols/diacylglycerols (yellow) was associated with higher risk (**Supplementary Table S8**).

## DISCUSSION

This prospective study of circulating pre-diagnostic metabolites provides evidence of an association between lipid metabolism and advanced prostate cancer risk. Several glycerolipids were suggestively positively associated with risk, particularly in lean individuals. The strongest classes identified in enrichment analyses were lipid-related: overall and unsaturated diacylglycerols, overall and unsaturated triacylglycerols, and acylcarnitines. Most metabolites were lipids of different chain lengths and fatty-acid desaturations,^19^ allowing for identification of novel lipid biomarkers of prostate cancer progression.

Our findings align with a case-only analysis in the Alpha-Tocopherol, Beta-Carotene Cancer Prevention Study that found several fatty acid lipid classes enriched for prostate cancer-specific mortality (197 cases, 92 deaths).^20^ They also align with experimental evidence highlighting the role of lipid metabolism, including lipid uptake, de novo synthesis, storage transport, and oxidation, in advanced prostate cancer development.^21^ Increased lipid production in a subset of prostate tumors – a ‘lipogenic phenotype’ – aids in metastatic spread and serves as an early marker of prostate cancer development and severity.^21^ Of 16 lipids nominally associated with advanced prostate cancer in our study, higher levels of 10 were associated with higher risk, suggesting that circulating lipids may indicate disease aggressiveness.

Diacylglycerols were most strongly associated with higher risk of advanced prostate cancer in enrichment analyses, and six diacylglycerols (C32:2, C36:1, C36:2, C36:3, C36:4, C38:4) showed nominal associations in at least one of the individual metabolite analyses (minimally-adjusted, BMI-adjusted, or cholesterol-adjusted). Diacylglycerols in cells act as secondary lipid messengers in the protein kinase C signaling pathway that can influence tumor progression and metastasis.^21, 22^ Mutations in diacylglycerol-regulating enzymes have been associated with prostate cancer aggressiveness and recurrence, as well as the transition from androgen-dependence to androgen-independence.^22, 23^ However, the relationship between circulating diacylglycerol levels and tumor cell signaling is unclear. Nevertheless, diacylglycerols C36:3 and C36:4 are part of the linoleic acid and derivative subclass,^24^ which may promote cell proliferation and prostate tumor growth.^25^ Diacylglycerols C32:2, C36:1, C36:2, and C38:4 are in the diradylglycerol subclass, whose role in prostate cancer remains unknown. More broadly, several diacylglycerols are impacted by diet quality^26^ and physical activity,^19^ both of which are associated with advanced prostate cancer.^27^ These diacylglycerols are transported to peripheral tissue as very low-density lipoprotein.^19^

We previously found that diacylglycerols C36:1, C36:2, C36:3, and C36:4 were positively associated with BMI, derived fat mass, and waist circumference in the HPFS.^12^ Our current study found that associations between these diacylglycerols and advanced prostate cancer were limited to lean men. Similarly, associations for other metabolites differed in lean versus overweight individuals. There are two potential interpretations of these findings. First, the signal for metabolites associated with advanced prostate cancer may be dampened by the diverse metabolic and hormonal pathways dysregulated by obesity. Second, because baseline advanced prostate cancer risk is higher with obesity,^2-5^ on the relative scale, ORs are smaller. While some prior studies have investigated associations of some metabolites with prostate cancer stratified by BMI,^20, 28-31^ none have reported associations for all metabolites or metabolite classes. Future studies are needed to replicate findings.

The yellow WGCNA module was comprised of 11 unsaturated diacylglycerols and 14 unsaturated triacylglycerols. Among the latter, eight were nominally associated with higher advanced prostate cancer risk in at least one individual metabolite analysis. As major components of very low-density lipoprotein and chylomicrons, triacylglycerols are energy sources and a means to transport fatty acids.^32^ To our knowledge, evidence for triacylglycerols and prostate cancer risk is limited.

This study also found acylcarnitines, including in one WGCNA module, to be associated with lower advanced prostate cancer risk. In the European Prospective Investigation into Cancer and Nutrition, higher acylcarnitine levels (C0, C14:1, C18:1, and C18:2) were associated with lower advanced prostate cancer risk.^33^ Decreased serum carnitine levels have been reported in numerous cancers, including endometrial and breast.^34^ Carnitine is a cofactor for transport of fatty acids across the mitochondrial membrane and thus plays a role in β oxidation of fatty acids as a potential source of ATP for cancer cells.^34^ *In vitro* studies of colon cancer cells have shown that low carnitine levels decreased mitochondrial fatty acid uptake.^35^ Exogenous carnitine facilitated increased fatty acid uptake, in turn increasing β-oxidation.^35^ *In vitro* studies of prostate cancer have shown that acetyl-L-carnitine reduces tumor cell migration and adhesion, potentially due to an anti-inflammatory role and suppression of pro-angiogenic factors.^36^

Lower grade advanced tumors showed enrichment for lipids and lipid metabolites, while higher grade advanced tumors were enriched for plasmalogens, acylcarnitine, and amines. Similarly, a small study of tumor tissue metabolites found higher long-chain acylcarnitines (palmitoylcarnitine and stearoylcarnitine) levels in Gleason score 9 relative to 7 tumors, potentially due to the role of acylcarnitines in fatty acid oxidation;^37^ lipid metabolite classes also differentiated high and low grade tumors. Separately, polyamines were the top-ranked pathway comparing Gleason score 8-10 to lower grade tumors (*p*_adjusted_=0.09).^38^ Future work should further characterize the relationships of lipids, acylcarnitines, and polyamines with higher Gleason to investigate the clinical utility of metabolomic lipid biomarkers in advanced disease detection.

Associations for lipids were strongest in cases diagnosed <5.5 years since blood draw, suggesting that circulating lipid levels could serve as an early detection biomarker. The amines and acylcarnitines associated with later cancers suggest a potential etiologic role in priming prostate carcinogenesis for developing aggressive tumors. Further investigations into these metabolite classes are warranted to determine if they are directly related to advanced disease development or represent separate underlying systemic metabolic processes that contribute to cancer progression.

A strength of our study was the prospectively collected blood linked to long-term follow-up and clinical annotation. While our study uniquely focused on advanced prostate cancer and is one of the largest of its kind, it was underpowered to detect statistically significant metabolites after multiple comparison adjustment. Nevertheless, we identified common lipid-related metabolic pathways dysregulated in advanced disease. Because fasting status can impact the levels of many metabolites, it would have been preferable for all participants to have fasted prior to blood draw. To address this limitation, we adjusted for fasting status in our models. The analyses stratified by post-baseline factors (i.e., time since blood draw and Gleason score) may have been subject to selection bias. We elected to include these analyses due to the importance of differentiating early detection from etiologic biomarkers for aggressive disease. In addition, given the long natural history of prostate cancer, we would not expect any bias to be substantial. Most participants, as well as those from other study populations that have investigated related research questions,^9^ were of European ancestry and may not capture some of the risk factors and social stressors of other populations. Given large racial disparities in prostate cancer outcomes^39^ and potential differences in metabolites among populations with different biological exposures and lived experiences,^40^ a key future direction is to undertake metabolomic studies in diverse populations.

In summary, this study identified associations between pre-diagnostic plasma lipids and advanced prostate cancer development, particularly in lean individuals and cases diagnosed <5.5 years from sample collection. It provides evidence of the potential clinical utility of lipids (notably diacylglycerols) as early detection biomarkers for advanced prostate cancer. Future work should replicate these findings in diverse cohorts and elucidate underlying biological mechanisms.

## Abbrevations

BMI: body mass index
CI: confidence interval
DAG: diacylglycerol
HPFS: Health Professionals Follow-up Study
OR: odds ratio
PSA: prostate-specific antigen
TAG: triacylglycerol
WGCNA: weighted correlation network analysis

## ACKNOWLEDGMENTS

We would like to thank the participants and staff of the HPFS for their valuable contributions as well as the following state cancer registries for their help: AL, AZ, AR, CA, CO, CT, DE, FL, GA, ID, IL, IN, IA, KY, LA, ME, MD, MA, MI, NE, NH, NJ, NY, NC, ND, OH, OK, OR, PA, RI, SC, TN, TX, VA, WA, and WY. The authors assume full responsibility for analyses and interpretation of these data. Where authors are identified as personnel of the International Agency for Research on Cancer / World Health Organization, the authors alone are responsible for the views expressed in this article and they do not necessarily represent the decisions, policy or views of the International Agency for Research on Cancer / World Health Organization.

## DATA AVAILABILITY STATEMENT

The metabolomic data used in this study have been deposited in the Harvard Dataverse (https://dataverse.harvard.edu/dataset.xhtml?persistentId=doi:10.7910/DVN/FR0CCH). R code is publicly available on GitHub (https://github.com/baileyvaselkiv/hpfs-prostate-cancer-metabolomics). Other data that support the findings of this study are available from the Channing Division of Network Medicine, but restrictions apply to the availability of these data. The data were used under license for the current study, and so are not publicly available. Data are, however, available from the authors upon reasonable request and with permission of the Channing Division of Network Medicine.

## ETHICS STATEMENT

The study protocol was approved by the institutional review boards of the Brigham and Women’s Hospital and Harvard T.H. Chan School of Public Health, and those of participating registries as required.

## AUTHORS’ CONTRIBUTIONS

LAM and PWK conceptualized the study design and oversaw its implementation. CBC and MGVH provided expertise on the metabolomics assays and interpretation. EME performed all analyses. REG, HF, BFD, and LAM completed initial interpretation of the results and prepared the first draft manuscript. All authors read and approved the final manuscript.

