## Supplementary Figures for "Pre-diagnostic lipid metabolites are enriched in men who develop advanced prostate cancer: a nested case-control study"

**Supplementary Figure S1.** Heatmap plot of the topological overlap matrix and cluster dendrogram for the 243 metabolites in this study. The colors on the axes represent the modules produced by WGCNA. The degree of metabolite overlap is represented by the shade of color; red represents higher co-expression connectivity and yellow represents lower co-expression connectivity.

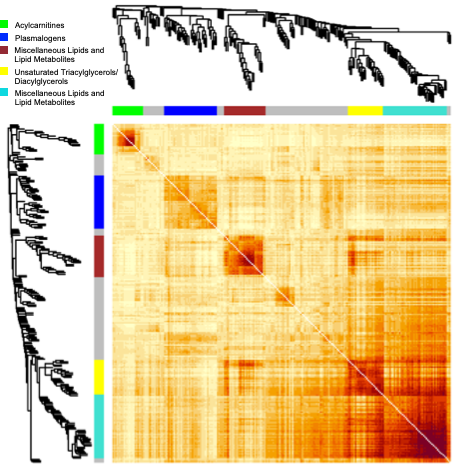
